# DeepSpot: Leveraging Spatial Context for Enhanced Spatial Transcriptomics Prediction from H&E Images

**DOI:** 10.1101/2025.02.09.25321567

**Authors:** Kalin Nonchev, Sebastian Dawo, Karina Silina, Holger Moch, Sonali Andani, Tumor Profiler Consortium, Viktor H Koelzer, Gunnar Rätsch

## Abstract

Spatial transcriptomics technology remains resource-intensive and unlikely to be routinely adopted for patient care soon. This hinders the development of novel precision medicine solutions and, more importantly, limits the translation of research findings to patient treatment. Here, we present DeepSpot, a deep-set neural network that leverages recent foundation models in pathology and spatial multi-level tissue context to effectively predict spatial transcriptomics from standard H&E images. DeepSpot substantially improved gene correlations across multiple datasets from patients with metastatic melanoma, kidney, lung, or colon cancers as compared to previous state-of-the-art. Using DeepSpot, we generated 3,780 TCGA virtual spatial transcriptomics samples (56 million spots) of the melanoma, renal cell cancer, lung adenocarcinoma and lung squamous cell carcinoma cohorts. We anticipate this to be a valuable resource for biological discovery and a benchmark for evaluating spatial transcriptomics models. We hope that DeepSpot and this dataset will stimulate further advancements in virtual spatial transcriptomics analysis.

## 1 Introduction

Spatial transcriptomics provides valuable insights into tissue-specific properties both in normal physiology [1–3] and in disease progression and treatment [4–8]. Despite its transformative potential, the technology remains technically demanding, resource-intensive, and currently lacks clinically validated prognostic or predictive biomarkers. Consequently, its integration into routine clinical practice remains limited [9, 10]. This constrains the development of new precision medicine solutions and, more importantly, limits the translation of basic research findings to patient diagnosis and treatment. To address these limitations, robust and cost-effective methods are urgently needed to broaden access and facilitate the generation of large-scale multimodal spatial transcriptomics datasets. Such resources will be critical for advancing cancer biology, enabling biologically informed tumor classifications, and laying the foundation for future clinical stratification and targeted therapies.

Recent advances in deep learning demonstrated that high-resolution hematoxylin and eosin (H&E)-stained slides can be used as input for computational models to efficiently predict bulk RNA expression [11–13]. Building on this, the increased availability of spatial transcriptomics data [5, 14–16] enabled the development of models to predict spatial transcriptomics from H&E images. However, accurately predicting spatial transcriptomics profiles remains challenging. Existing frameworks either struggle to effectively leverage morphological details and spatial context - especially those adapted from bulk RNA predictions - are computationally expensive or are limited to predicting only a small number of genes. For example, ST-Net uses a convolutional neural network (DenseNet-121 [17], pre-trained on ImageNet [18]) followed by a fully connected linear layer to predict the expression of 250 genes [19]. While Jaume and Doucet et al. [14] utilized recent pathology foundation models (e.g., UNI [20], Phikon [21]), pre-trained on extensive H&E datasets, to extract tile spot features, they focused on predicting only 50 genes using either ridge regression or random forest. Other methods, such as BLEEP [22], use contrastive learning to create a low-dimensional joint embedding and require access to the full training data during inference to perform k-nearest neighbor mapping. Furthermore, alternative methods employing vision transformers propose compressing the H&E image as a sequence of tiles and integrating their spatial location by encoding absolute pixel coordinates through positional embeddings to predict ∼750 genes [23–25]. However, the gigapixel resolution of pathology slides makes full-image processing impractical, requiring tile subsampling and potentially leading to the loss of important details. By contrast, the spatial tissue context, proven effective for representing spatial transcriptomics data [5], remains underutilized for predicting spatial transcriptomics from H&E images. Therefore, a new and reliable method is needed to overcome the aforementioned challenges, advance spatial transcriptomics prediction and enable its effective integration into clinical practice.

To address this, we developed DeepSpot, a novel deep-learning model that utilizes recent pathology foundation models and spatial multi-level tissue context to effectively predict spatial transcriptomics from H&E images. We compared its performance against previous state-of-the-art methods across multiple spatial transcriptomics datasets, including both spot-level (10x Genomics™, Visium) and single-cell-level (10x Genomics™, Xenium) technologies: 36 Visium and 20 Xenium lung cancer samples [26, 27], 24 Visium kidney cancer samples [28], 18 Visium metastatic melanoma samples [5], 8 Visium colon cancer samples [29], and 6 Visium samples enriched with tertiary lymphoid structures (TLS) from kidney and lung cancer. We demonstrated substantially improved gene correlations which enabled downstream virtual spatial transcriptomics analysis leading to biomarker and pathway discovery. Furthermore, we applied DeepSpot to 3,780 slide images from TCGA patients with corresponding bulk RNA-seq. This not only served as an out-of-distribution validation but also generated a large multimodal virtual spatial transcriptomics dataset with over 56 million spots from patients with melanoma, kidney or lung cancer. This dataset represents a unique resource that significantly enriches the available spatial transcriptomics data for TCGA samples, providing valuable insights into the molecular landscapes of cancer tissues (Figure 1A).

**Fig 1.**
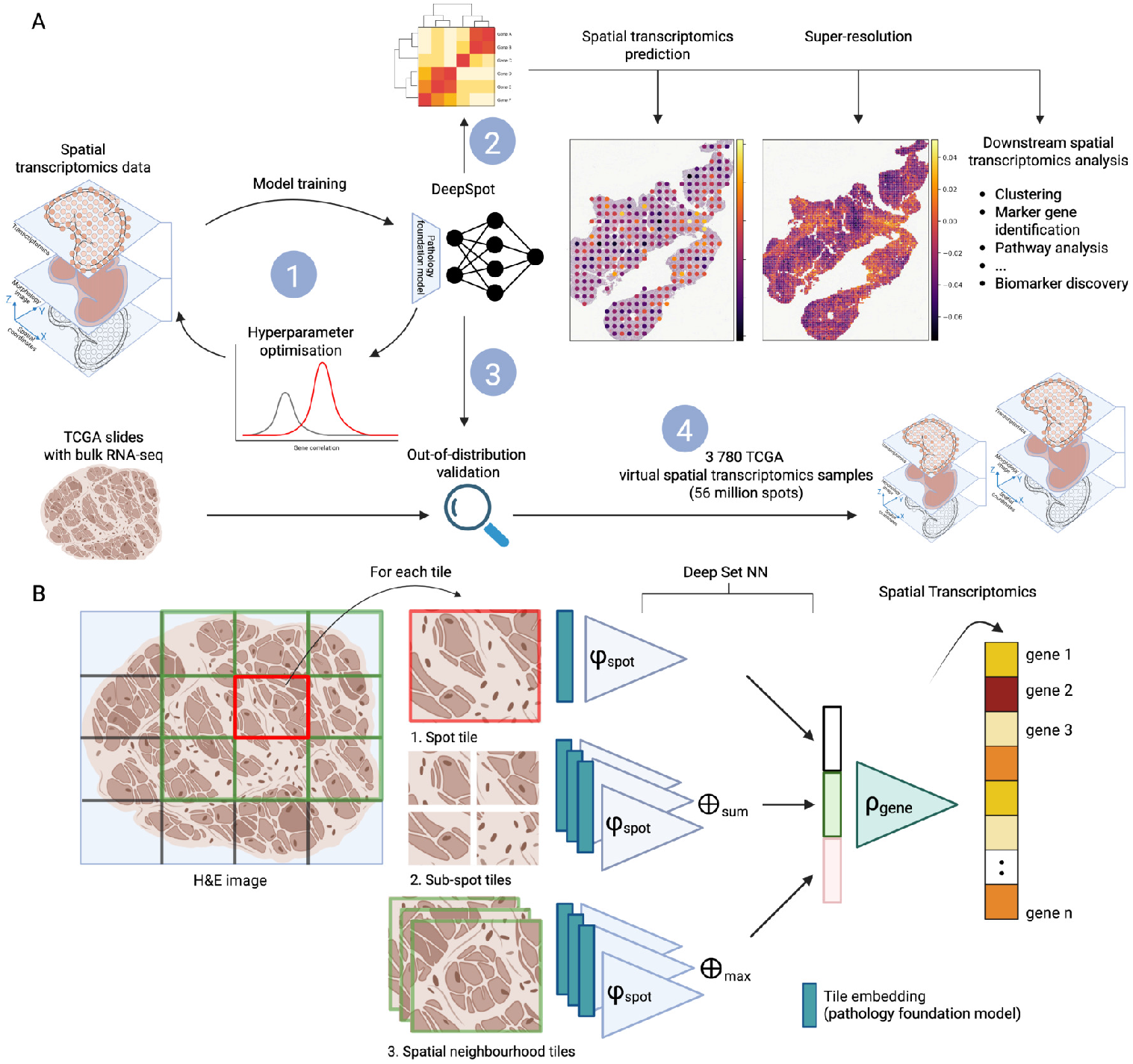
DeepSpot leverages pathology foundation models and spatial tissue context. **A:** Graphical summary. DeepSpot predicts spatial transcriptomics from H&E images by leveraging recent foundation models in pathology and spatial multi-level tissue context. **1:** DeepSpot is trained to predict 5,000 genes, with hyperparameters optimized using cross-validation. **2:** DeepSpot can be used for de novo virtual spatial transcriptomics prediction or for correcting existing spatial transcriptomics data. **3**: Outof-distribution validation is performed using nested leave-one-out cross-validation at the patient level and independent testing on external cohorts. We predicted spatial transcriptomics from TCGA slide images, aggregated the data into pseudo-bulk RNA profiles, and compared them with the available ground truth bulk RNA-seq. **4**: DeepSpot generated 3,780 TCGA virtual spatial transcriptomics samples with over 56 million spots from melanoma, kidney or lung cancer patients, enriching the available spatial transcriptomics data for TCGA samples and providing valuable insights into the molecular landscapes of cancer tissues. **B**: Workflow of DeepSpot. H&E slides are first divided into tiles, each corresponding to a spot. For each spot, we create a bag of sub-spots by dividing it into sub-tiles that capture the local morphology and a bag of neighboring spots to represent the global context. A pre-trained pathology foundation model extracts tile features, which are input to the model. The concatenated representations are then fed into the gene head predictor, ρ_*gene*_, to predict spatial gene expression.

## 2 Results

### 2.1 DeepSpot leverages pathology foundation models and spatial tissue context

We developed DeepSpot, a novel deep-learning model that leverages recent foundation models in pathology to effectively predict spatial transcriptomics from H&E images. DeepSpot employs a deep-set neural network [30] to model transcriptomic spots as bags of sub-spots and integrates multi-level tissue details along with spatial neighborhood morphology. This integration, supported by the robust foundation from pre-trained H&E models, significantly enhances the accuracy and granularity of gene predictions from H&E images at minimal extra cost (Figure 1B). The model is trained to predict the 5,000 most variable genes in a multiregression setting using spatial transcriptomics data. First, the H&E slides are split into tiles, each corresponding to a transcriptomic spot. For each spot, we create a bag of sub-spots by splitting it into non-overlapping sub-tiles to capture local tissue morphology and a bag of neighboring spots to capture the global tissue environment. A pre-trained pathology foundation model (e.g., UNI [20], Phikon [21], H-optimus-0 [31]) is then used to extract tile feature representations. These features input DeepSpot’s *φ*_*spot*_ module, whose learned representations are concatenated and fed into the gene head predictor, *ρ*_*gene*_, to predict the spatial transcriptomics profiles. Ultimately, the predicted virtual spatial transcriptomics can be used for various downstream tasks, including spot phenotyping, gene expression analysis, and pathway and biomarker discovery.

The motivation for modeling transcriptomic spots as bags of sub-spots is inspired by the fact that each spot contains between 1-10 cells (10x Genomics™, Visium). This approach captures finer details within the target spot, effectively overcoming the resolution limitations of the sequencing technology while also learning the contributions of individual sub-spots. Additionally, DeepSpot integrates the global tissue environment by pooling neighboring spots and jointly learning the tissue landscape. Building on the foundation of recent models for H&E images, the deep sets architecture is particularly well-suited for representing spatial transcriptomics data due to its permutation invariance and permutation equivariance properties [30]. These characteristics make it an effective framework for modeling spatial transcriptomics, as it effectively captures the relationships between spots by focusing on biologically meaningful spatial patterns rather than the exact order of the spots (Figure S1). Furthermore, the shared weights of the *φ*_*spot*_ module are optimized simultaneously across multiple tasks, leading to better generalization and more efficient learning in the limited data context of spatial transcriptomics data.

### 2.2 DeepSpot improves spatial gene expression prediction

We benchmarked DeepSpot’s performance on multiple spatial transcriptomics datasets sequenced using Visium from 10x Genomics™ to predict the top 5,000 most variable genes as defined by Scanpy with the Seurat v3 flavor [34]. In line with the methodology of [14, 19, 22–25], we adopted Pearson correlation to measure the similarity between the predicted and the ground truth gene expression. We calculated correlations for the top 50 to 5,000 most predictive genes (Figure 2A). To avoid hyperparameter overfitting, we employed nested leave-one-out patient cross-validation, with an internal cross-validation loop for hyperparameter selection. Specifically, for each fold, we trained a model on the training set, selected hyperparameters on the validation set, and evaluated performance on the test set, ensuring that each set contains distinct patients. This process was iterated over all folds, and the resulting median Pearson correlation, along with the standard error, is reported (Figure 2A, S1).

**Fig 2.**
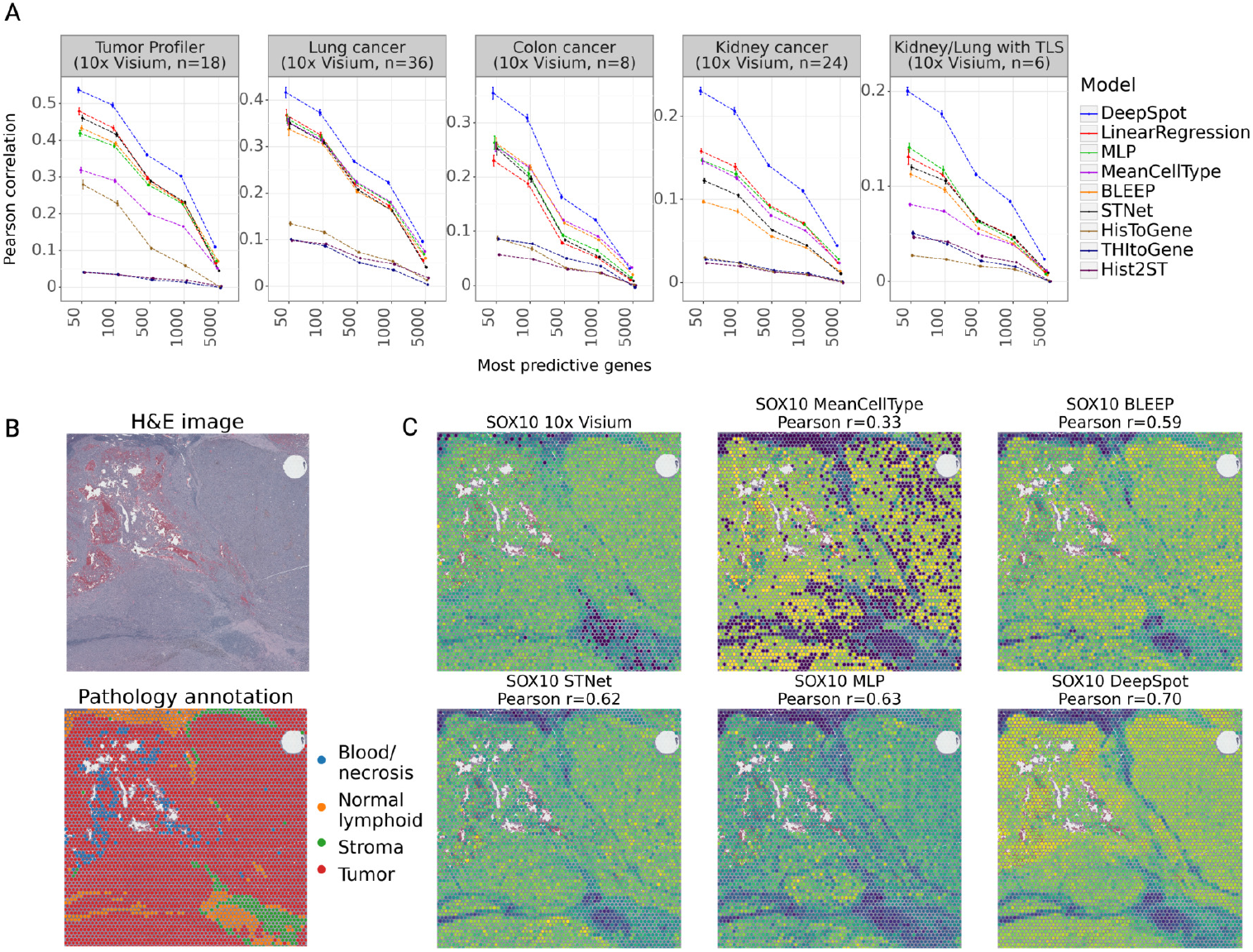
DeepSpot improves spatial gene expression prediction. **A**: Benchmark of DeepSpot and previous methods on five spatial transcriptomics datasets generated using Visium from 10x Genomics™. The y-axis represents the average Pearson correlation between the predicted and ground truth gene expression. The x-axis represents Pearson correlation computed on the top N predictive genes per model. Models are ordered based on their relative rank across datasets. **B**: H&E image and pathology annotation for slice MELIPIT-1-1 from the Tumor Profiler dataset and **C**: Comparison of gene expression for *SOX10* (melanoma marker [32, 33]).

DeepSpot consistently predicted improved spatial gene expression with higher Pearson correlation across all datasets compared to other methods (Figure 2A, S2, S3). For example, on the Tumor Profiler dataset [5], DeepSpot improved the Pearson correlation across the top 1000 genes by 30%, increasing it from 0.23 (best competitor, STNet) to 0.30 (DeepSpot), with some genes exceeding a correlation of 0.60 (Figure S3). The Tumor Profiler dataset comprises 18 tissue slices from 7 patients with metastatic melanoma.

This improvement highlights the superior performance of DeepSpot in predicting spatial transcriptomics by leveraging recent foundation models in pathology and spatial multi-level tissue context. The vision transformer-based models (HisToGene, Hist2ST, and THItoGene) underperformed across all datasets, likely because their architecture depends on learning pathology features from scratch. In contrast, leveraging transfer learning with prior histopathology knowledge from pathology foundation models trained on extensive collections of clinical slides could offer a significant advantage (Table S2, Figure S1).

To qualitatively illustrate the predicted virtual spatial transcriptomics, we compare the expression for *SOX10*, a known melanoma marker gene [32, 33] for slice MELIPIT-1-1 with available ground truth pathology annotations (Figure 2B, C).

As a baseline, predicting the mean cell type gene profile (MeanCellType, *ρ* = 0.33) captures only broad expression patterns but lacks spatial and tumor-specific details. Gene expression predictions from BLEEP (*ρ* = 0.59) and ST-Net (*ρ* = 0.62) exhibit low spatial resolution, with areas of high expression intensity concentrated at tumor boundaries. In contrast, baseline MLP (*ρ* = 0.63), utilizing pathology foundation model features, shows greater contrast between normal and tumor tissues, suggesting improved discrimination. However, the expression patterns within tumor regions remain noisy and spatially disorganised, limiting the interpretability of the tumor microenvironment. Notably, DeepSpot achieves the highest gene correlation to the ground truth (*ρ* = 0.70), effectively distinguishing between normal and tumor tissues. DeepSpot produced a spatially organized gene expression pattern within the tumor, with the highest levels of expression concentrated in the tumor’s core (Figure 2C).

Overall, an in-depth gene ontology enrichment analysis of the top predicted genes indicates that DeepSpot effectively learns genes involved in key cancer progression processes, including signaling, immune response, proliferation, apoptosis, adhesion, and extracellular matrix organization [35–38](Figure S4). In contrast, genes with lower prediction scores were associated with broader, less specific pathways (e.g., population dynamics, positive regulation), reflecting generalized rather than specific biological processes.

### 2.3 DeepSpot enables virtual spatial transcriptomics analysis

To demonstrate the utility of the virtual spatial transcriptomics data, we use the DeepSpot predicted gene expression for slice MELIPIT-1-1 from the Tumor Profiler dataset to perform a common spatial transcriptomics analysis workflow [39]. First, we apply dimensionality reduction to visualize the data with randomly sampled morphology spots (Figure 3A). We observe an aggregation of tumor spots on the right side, with a gradual transition towards spots of necrosis, stroma, and normal lymphoid tissue on the left. Tumor spots appear significantly larger with enlarged nuclei; stroma cells have an elongated morphology and are noticeably more scattered; normal lymphoid cells are generally smaller and denser in line with the observations in [5].

**Fig 3.**
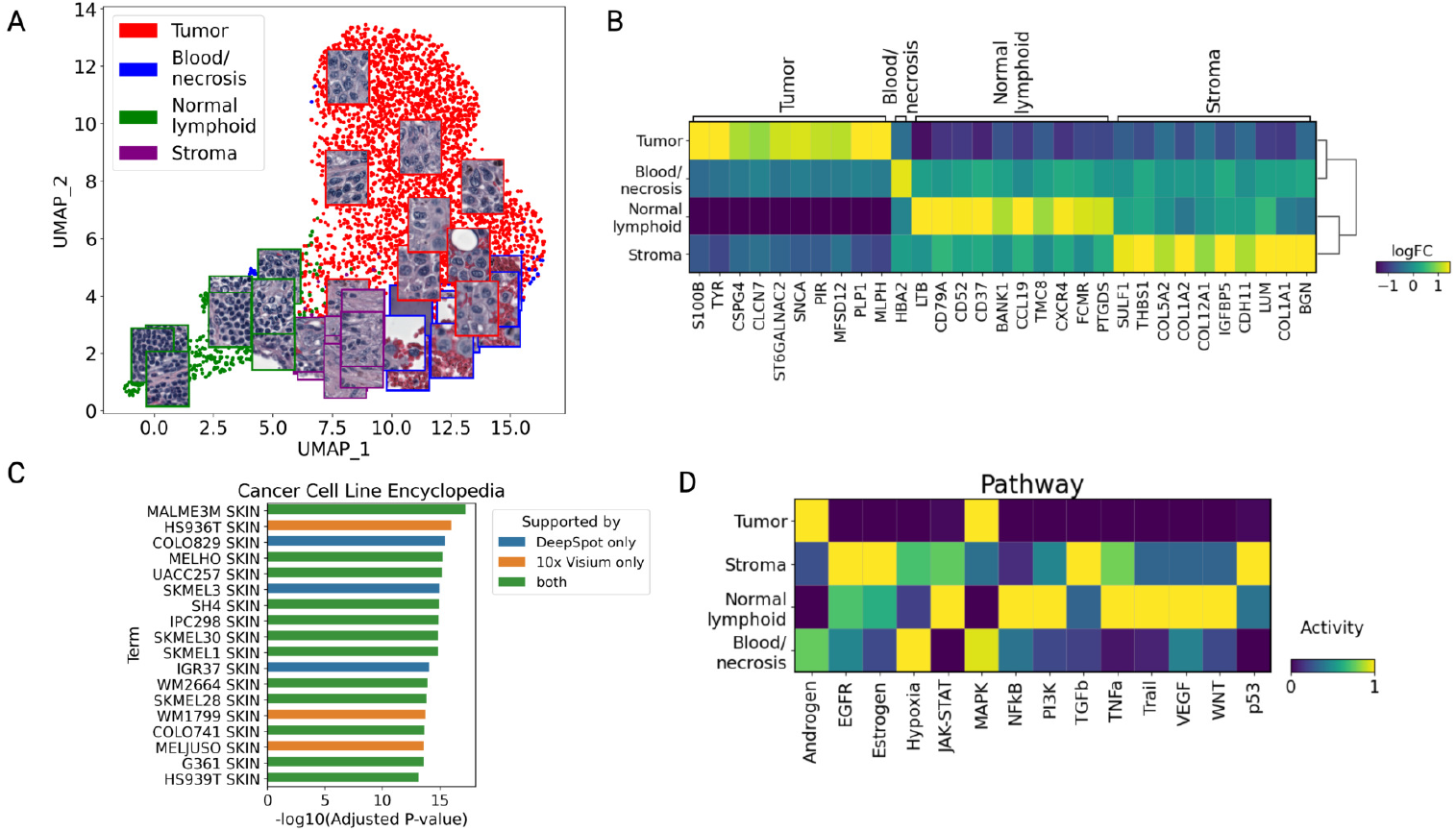
DeepSpot enables virtual spatial transcriptomics analysis. **A**: UMAP plot of virtual spatial transcriptomics from DeepSpot for sample MELIPIT-1-1 from the Tumor Profiler dataset with sampled spot images. **B**: Matrix plot of the log fold change (logFC) for significant marker genes (logFC > 1) per group, derived from virtual spatial transcriptomics of the same sample. **C**: Gene set enrichment analysis using the top 100 tumor marker genes and the data from the Cancer Cell Line Encyclopedia [45]. **D**: Pathway analysis based on the virtual spatial transcriptomics using decoupleR [51].

Next, we performed a marker gene analysis on the virtual spatial transcriptomics using the Wilcoxon test. Among the identified tumor marker genes, several well-known melanoma-related genes (e.g., *S100B* [40], *TYR* [38], *CSPG4* [41, 42], *PMEL* [43] and *MLPH* [44]) were found (Figure 3B) and subsequently validated with the ground truth spatial transcriptomics (Figures S5, S6). They are involved in melanoma progression and associated with increased invasiveness of melanoma cells [35–38]. To further support our findings, we performed enrichment analysis using the top 100 tumor marker genes in conjunction with data from the Cancer Cell Line Encyclopedia [45] (Figure 3C). The most enriched cell lines were related to skin melanoma, indicating a strong association with tumor biology relevant to these malignancies [46–50]. Comparison with the ground truth spatial transcriptomics analysis revealed an overlap of 80% in identified cell lines (Figure 3C, S7), further validating the robustness of our approach. Moreover, the significant enrichment of pathways related to cell proliferation and survival highlights potential therapeutic targets warranting further investigation.

Finally, we conducted a pathway analysis (Figure 3D) using decoupleR [51] which revealed increased activity of the MAPK pathway in tumor tissues. This pathway is known for promoting processes such as cell proliferation, invasion, metastasis, migration, survival, and angiogenesis [52–54]. Consistent with findings from the ground truth spatial transcriptomics (Pearson r = 0.74, Figure S8), we observed predominant hypoxia signatures in regions of necrosis and hemorrhage, while JAK-STAT inflammatory signaling was dominant within the cancer microenvironment. Overall, these transcriptomics analyses highlight the accurate gene expression predictions made by DeepSpot and reinforce their interpretation in the context of underlying biological processes.

### 2.4 DeepSpot overcomes spatial transcriptomics gene detection sensitivity

Higher resolution in spatial transcriptomics such as that produced by Visium from 10x Genomics often leads to lower gene sensitivity and spot contamination - known issues that result in poor downstream performance and misleading results [55]. These challenges are further compounded by the reduced RNA quality in archival samples. To demonstrate how DeepSpot addresses these issues, we present tissue slice KC2 (Figure 4A) excluded from the experiments due to low transcriptomics quality (Figure S9). Specifically, regions annotated as tertiary lymphoid structures (TLS) did not show expression of known TLS marker genes (Figure 4B, red boxes) [56, 57], and unexpectedly, these genes were detected in other locations. This hold-out sample would typically have been discarded from downstream analysis. However, DeepSpot, trained on kidney and lung samples with TLS, successfully recovered this low-quality sample by effectively transferring tissue morphology patterns associated with TLS. As a result, DeepSpot accurately predicted the gene expression at positions corresponding to these morphological features (Figure 4C). Moreover, DeepSpot’s architecture enables gene expression prediction at super-resolution by focusing on a single sub-spot at the center of the spot, further augmenting patient data with fine-grained details at minimal additional cost (Figure 4D). For example, we predicted spatial transcriptomics for the same patient by reducing spot distance (4x) and diameter (3x) in pixels, which enhanced data resolution and improved spatial localization. The superresolution allowed us to pinpoint the exact locations in the H&E image corresponding to the highest gene expression (e.g., *LTB*, Figure 4D). Specifically, one can observe that the highest *LTB* expression occurs in the center of the TLS formations, potentially indicating the TLS germinal centers.

**Fig 4.**
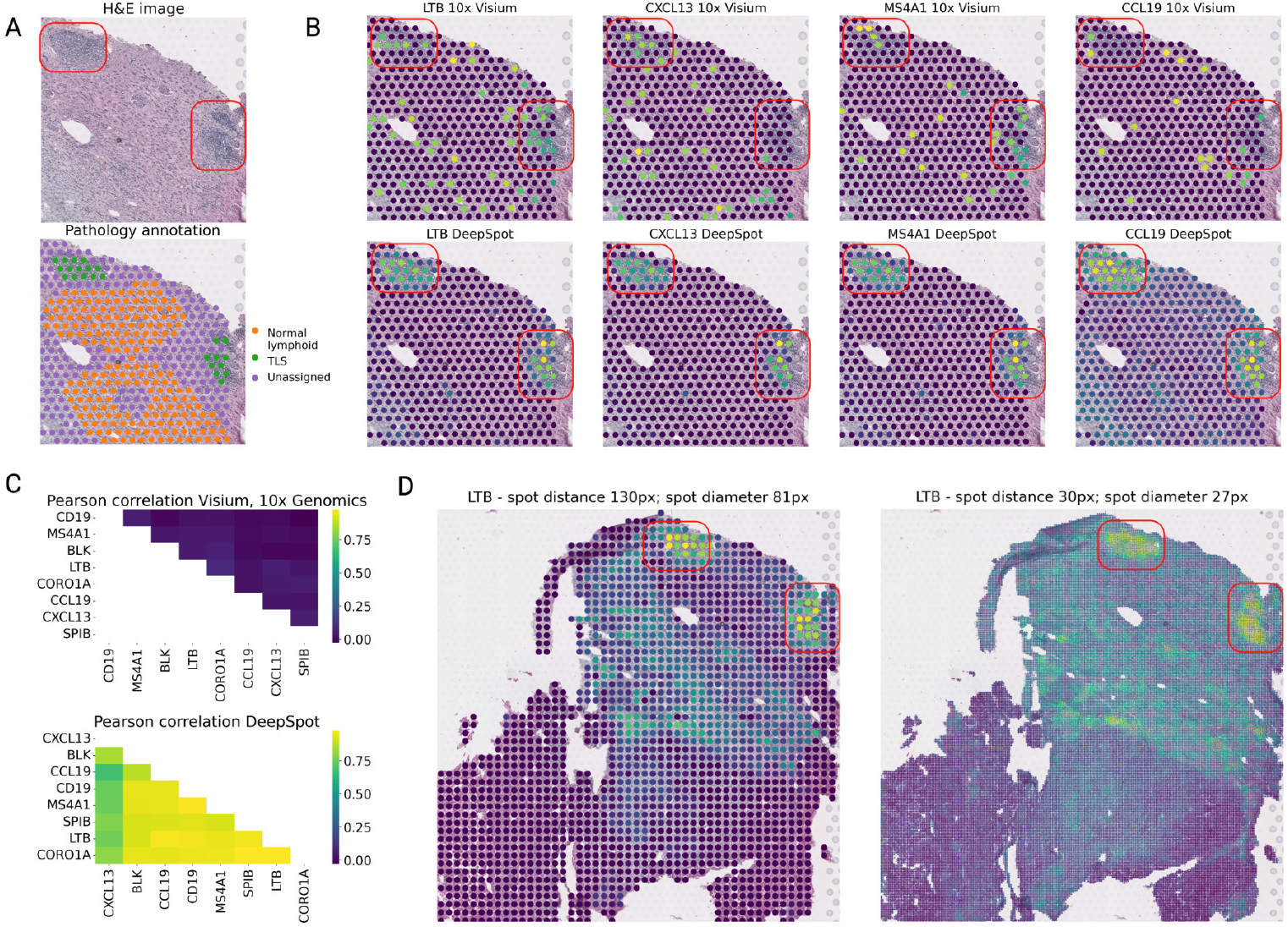
DeepSpot overcomes spatial transcriptomics gene detection sensitivity. **A**: H&E image and pathology annotation for slice KC2 from the Lung/Kidney with TLS dataset, zoomed into TLS formations. **B**: Comparison of gene expression for *LTB, CXCL13, MS4A1M*, and *CCL19* (known TLS markers) between ground truth data from 10x Genomics™, Visium and predictions from DeepSpot. **C**: Pearson correlation of gene expression for *LTB, CXCL13, MS4A1M, CCL19, CORO1A, CD19, SPIB*, and *BLK* across all spots in slice KC2. **D**: Spatial transcriptomics predictions for gene *LTB* with original spot distances of 130 px and spot diameter of 81 px and at super-resolution with spot distances of 30 px and spot diameter of 27 px in slice KC2.

### 2.5 DeepSpot predicts virtual spatial transcriptomics for 3,780 TCGA slides

To assess out-of-distribution performance, we validated DeepSpot, trained on melanoma, kidney or lung cancer spatial transcriptomics, using fresh frozen (FF) or formalin-fixed paraffin-embedded (FFPE) slides from 3,780 TCGA samples with corresponding bulk RNA-seq (Figure 5A). In summary, we predicted spatial gene expression for each slide in TCGA with melanoma (SKCM), renal cell (KIRC), lung adenocarcinoma (LUAD) or lung squamous cell carcinoma (LUSC), and then aggregated the expression to generate pseudo-bulk RNA profiles. We compared this pseudo-bulk RNA to the available ground truth bulk RNA-seq (Figure 5B, S10). Notably, DeepSpot accurately predicted gene expression and outperformed other models also in an out-of-distribution setting. Overall, the DeepSpot pseudo-bulk RNA profile derived from lower-quality FF slides correlated more closely with the paired bulk RNA from the same tissue piece compared to the larger FFPE slide. Although DeepSpot was trained on FFPE slides, its design, leveraging a pathology foundation model, enabled effective generalization to FF slides, highlighting its potential for reliable gene expression predictions (Figure 5B). Moreover, DeepSpot’s predictions were more accurate for genes with greater variability in the training data (Figure S10). This underscores the need for larger spatial transcriptomics datasets that encompass a diverse range of genes to train more precise models.

**Fig 5.**
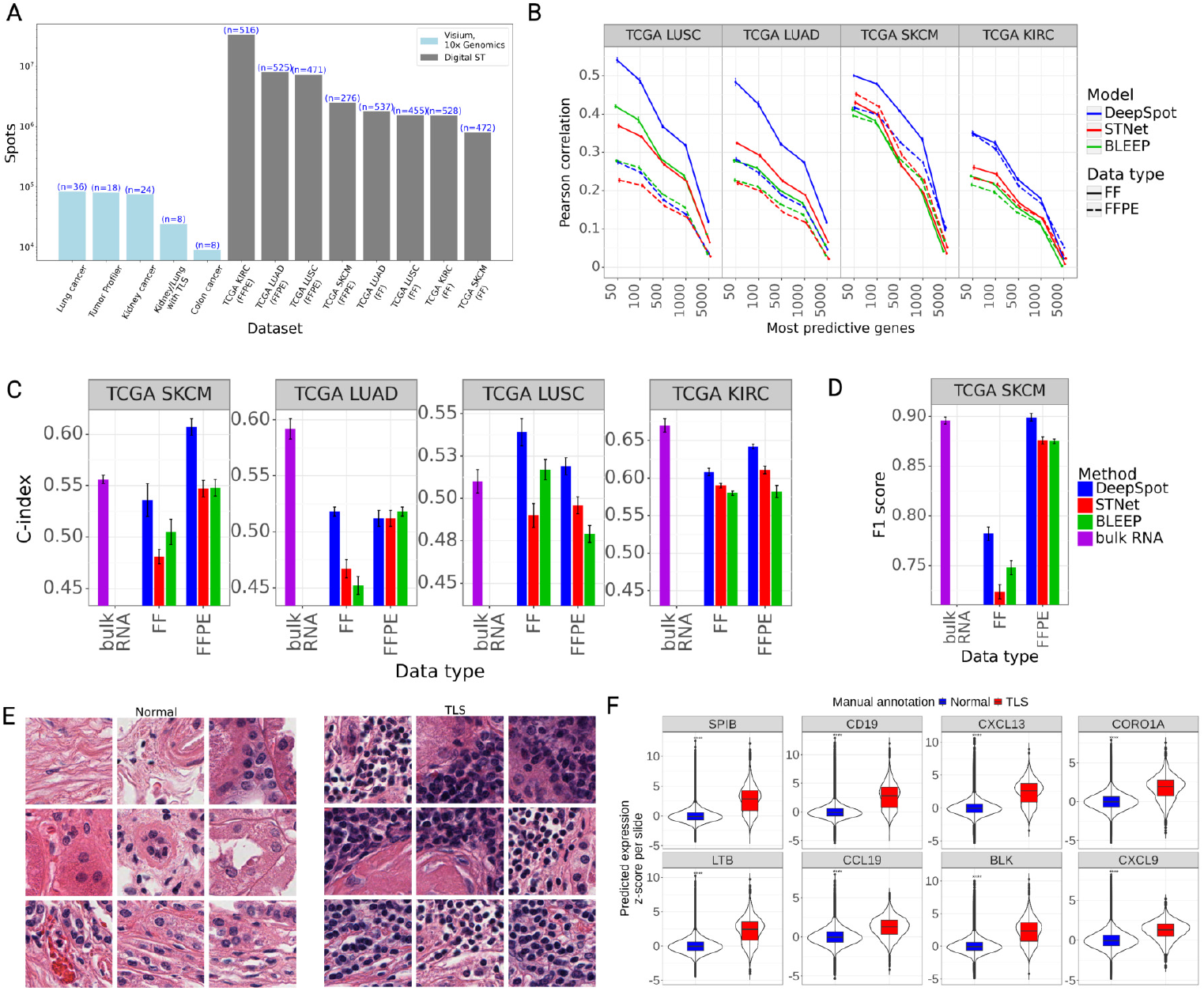
DeepSpot predicts virtual spatial transcriptomics from 3,780 TCGA slides. **A**: Number of spots and samples per dataset. The x-axis represents the datasets, while the y-axis shows the number of spots. The number of samples is displayed above each dataset bar. **B**: Out-of-distribution performance on models trained on 10x Genomics™, Visium and gene expression predictions based on TCGA slides. The y-axis represents the average Pearson correlation between the pseudo-bulk RNA and the ground truth bulk RNA profile. The x-axis represents Pearson correlation computed on the top N predictive genes per model. The line type indicates the data type used to generate the predictions (FF or FFPE). **C**: Survival analysis benchmark. The x-axis represents the concordance index (C-index), while the y-axis represents the data type. **D**: Tumor type classification benchmark. The x-axis represents the F1 score, while the y-axis represents the slide type. **E**: TCGA FFPE KIRC examples of spots manually labeled as normal tissue and TLS. **F**: The DeepSpot-predicted expression of known TLS marker genes in spots manually labeled as Normal or TLS. The y-axis represents the predicted gene expression, standardized per slide. Stars denote the levels of statistical significance, with **** indicating p<0.0001 as determined by the Wilcoxon rank-sum test.

To further demonstrate the quality and robustness of our virtual spatial transcriptomics data, we trained downstream models on the pseudo-bulk RNA for several tasks, including survival analysis (Figure 5C) and tumor type classification (Figure 5D). Notably, the gene expression generated from DeepSpot outperformed previous methods in survival analysis and in some instances, provided better prognostic power than the ground truth bulk RNA-seq data (Figure 5C). For instance, in the SKCM dataset, DeepSpot improved the concordance index by 11%, increasing it from 0.548±0.003 (best competitor, BLEEP FFPE) to 0.608±0.008 (DeepSpot FFPE). When compared to the ground truth bulk RNA-seq (0.556±0.004), this represents a 10% improvement. Moreover, DeepSpot’s gene expression improves tumor type classification in SKCM, increasing the F1 score by 3%, from 0.876±0.003 (best competitor, STNet FFPE) to 0.899±0.003 (DeepSpot FFPE), slightly surpassing the ground truth bulk RNA-seq - 0.895±0.003 (Figure 5D). Finally, we compared the DeepSpot-predicted gene expression of known TLS marker genes [56, 57] in FFPE KIRC samples with available manual annotations [58] (Normal tissue or TLS, Figure 5E). Notably, TLS marker genes were significantly overexpressed in spots annotated as TLS compared to those in normal tissue (Figure 5F), underscoring the robustness of DeepSpot-predicted spot-level gene expression in providing enhanced contextual insights.

These experiments resulted in a large multimodal virtual spatial transcriptomics resource with over 56 million spots from 3,780 samples with melanoma, kidney or lung cancer. It represents a unique resource that significantly enriches the available spatial transcriptomics data for TCGA samples, providing unique insights into the molecular landscapes of cancer tissues. To further illustrate the richness and utility of our dataset compared to the limited information from TCGA bulk RNA-seq, we compared the expression of the melanoma marker *SOX10* in both bulk RNA data (Figure S11) and virtual spatial transcriptomics data (Figure S12) for SKCM. *SOX10* expression is notably higher on the left side, indicating the presence of tumor spots, and gradually decreases towards the right. Additionally, we provide visual examples of predicted spots expressing *SOX10* (melanoma), *CD37* (normal lymphoid), and *COL1A1* (stroma) (Figure S13), which further validates the correlation between the morphological data and predicted transcriptomics. DeepSpot demonstrated to be a robust method for predicting spatial transcriptomics from H&E images, with the ability to generalize effectively to previously unseen and out-of-distribution pathology slides.

### 2.6 DeepSpot predicts single-cell spatial transcriptomics from H&E images

Spatial transcriptomics technology is evolving rapidly, with newer methods achieving single-cell resolution [59, 60]. To position DeepSpot within this advancing field, we developed an adapted version of DeepSpot for predicting single-cell spatial transcriptomics from H&E images. It modifies the architecture by removing the subspot module, enabling it to predict spatial transcriptomics from H&E images at the cellular level. To demonstrate its performance, we evaluated it on a publicly available single-cell lung cancer spatial transcriptomics dataset sequenced with Xenium from 10x Genomics™ [27]. DeepSpot improved the Pearson correlation across the top 100 genes by 81%, increasing it from 0.16 (best competitor, STNet) to 0.29 (DeepSpot) (Figure 6A). Moreover, the larger cohort size in this dataset facilitated a systematic assessment of model performance relative to training data size. DeepSpot’s accuracy improved consistently with increasing data size, indicating potential for further enhancement as additional spatial transcriptomics datasets become available (Figure 6B). Conversely, BLEEP’s performance declined with more training data, likely due to the increasing difficulty of accurately identifying nearest matching spots, resulting in less precise matches and diminished overall performance.

**Fig 6.**
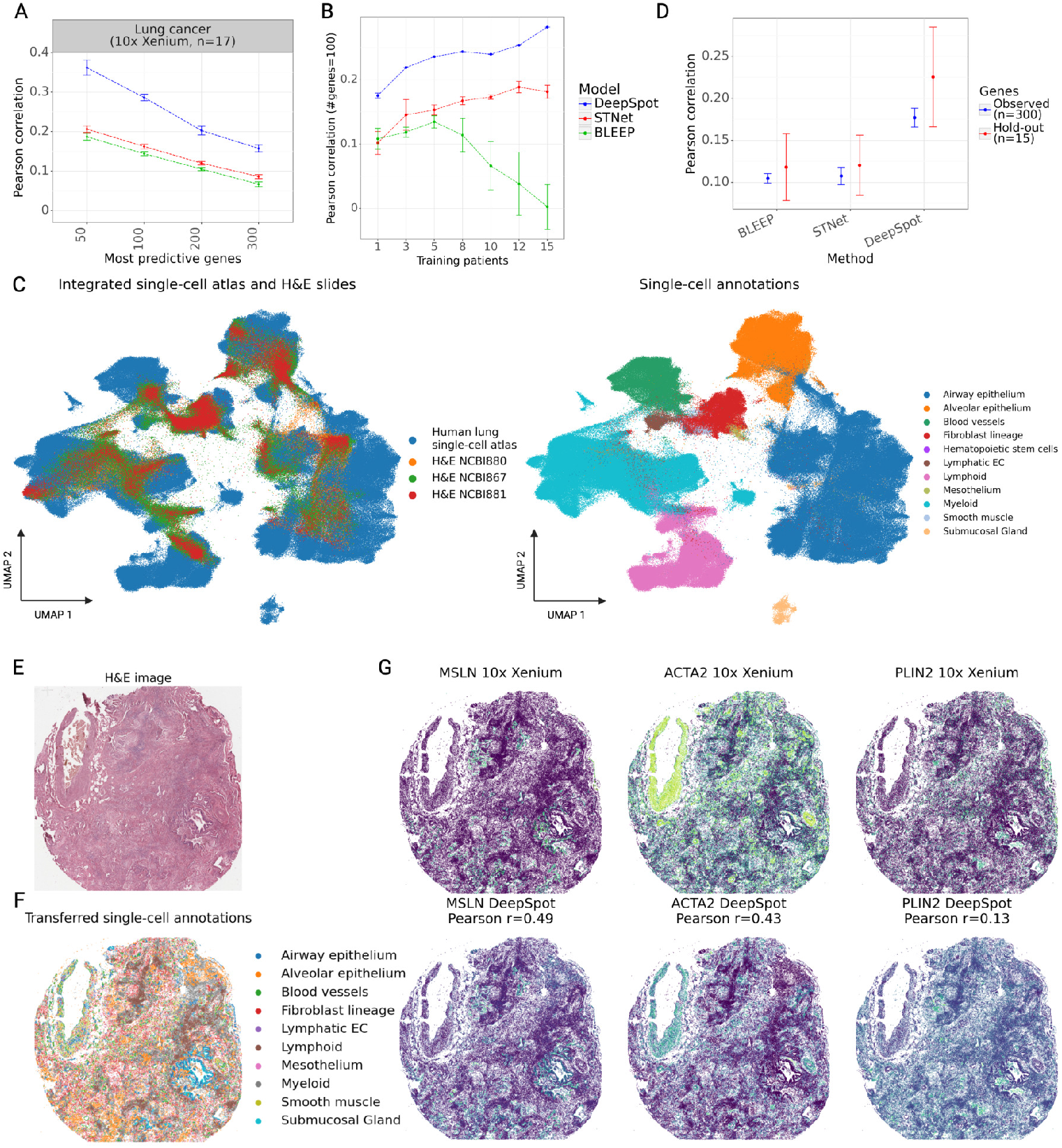
DeepSpot bridges H&E images and single-cell atlases at cellular resolution. **A**: Benchmark of DeepSpot and other methods on a single-cell spatial transcriptomics dataset generated using Xenium from 10x Genomics™. The y-axis represents the average Pearson correlation between the predicted and ground truth gene expression. The x-axis represents Pearson correlation computed on the top N predictive genes per model. **B**: Model performance as a function of training set size. The y-axis indicates the average Pearson correlation computed across the top 100 most variable genes. The x-axis represents the number of patients used for training. **C**: Integrated UMAP of the predicted single-cell spatial transcriptomics from H&E images with DeepSpot and a human lung single-cell reference atlas [62]. **D**: Measuring the gene expression correlation between the ground truth single-cell spatial transcriptomics data and the gene expression that resulted from mapping the virtual spatial transcriptomics onto the reference atlas. Observed genes (n = 300, blue) were used for training, while hold-out genes (n = 15, red) were withheld to assess generalization to unseen gene expression. The y-axis represents the Pearson correlation between the predicted and ground truth gene expression. **E**: H&E image for slice NCBI867 from the Xenium lung dataset **F**: Transferred annotations for the same slide from the single-cell lung reference atlas. **G**: Comparison of gene expression for the hold-out genes - *MSLN, PLIN2*, and *ACTA2* across ground truth data from 10x Genomics™, Xenium and the gene expression derived from mapping DeepSpot-predicted virtual spatial transcriptomics onto the reference atlas.

### 2.7 DeepSpot enables H&E image integration with single-cell atlases at cellular resolution

Due to the limited gene coverage provided by Xenium from 10x Genomics™, spatial transcriptomics data is typically aligned with a single-cell reference atlas to transfer knowledge (e.g., missing gene expression, annotations) [3, 7, 61]. To address this, we applied the pretrained single-cell models to three hold-out H&E slides from the Xenium lung dataset and inferred the predicted virtual singlecell spatial transcriptomic profiles for the 300 genes used during training. We then mapped the predicted gene expression onto an annotated lung single-cell reference atlas [62] comprising 584,944 cells and 27,957 genes, using the shared genes as common coordinates for alignment (Figure 6C).

We measured the Pearson correlation between the ground truth single-cell spatial transcriptomics data and the gene expression inferred through the mapping onto the reference atlas. This was done for the 300 genes used during training, as well as 15 genes that we hold out to assess generalization to unseen gene expression (Figure 6D, Table S2). By exploiting gene co-expression patterns, DeepSpot significantly outperformed previous models and improved the Pearson correlation across the holdout genes by 64%, increasing it from 0.14 (best competitor, STNet) to 0.23 (DeepSpot) (Figure 6D). This aproach enabled us to transfer information from the large well-curated lung single-cell reference atlas onto new unlabeled H&E images (Figure 6E) by either assigning cell annotations (Figure 6F) or spatially inferring the expression of unobserved genes (Figure 6G). These results emphasize the ability of our model to integrate two traditionally independent modalities - H&E images and transcriptomic single-cell reference atlases, by aligning them in a common space, enabling efficient knowledge transfer. This facilitates the inference of spatial gene expression patterns from H&E images and offers a more comprehensive understanding of tissue architecture.

## 3 Discussion

In this work, we propose DeepSpot, a method that leverages pathology foundation models and spatial tissue context to effectively predict spatial transcriptomics from routine histology images. Our results consistently show improved spatial gene predictions over previous methods across multiple datasets, including melanoma, colon, kidney and lung cancers sequenced using spot-level (10x Genomics™, Visium) and single-cell-level (10x Genomics™, Xenium) technologies. Importantly, DeepSpot demonstrated robustness to out-of-distribution H&E images from TCGA, producing accurate gene predictions. This led to the creation of a unique multimodal virtual spatial transcriptomics dataset with over 56 million spots from 3,780 samples with melanoma, kidney or lung cancer. We systematically demonstrated that the gene expression obtained from DeepSpot enables de novo virtual spatial transcriptomics analysis, including clustering, marker gene identification, pathway discovery and single-cell reference atlas integration.

This improvement in spatial transcriptomics prediction resulted from incorporating multi-level tissue details and spatial neighborhood information along with utilizing pathology foundation models trained on extensive datasets of H&E slides. Our approach addresses the limitations of sequencing technology resolution by representing spots as bags, aggregating local spot information, and pooling global tissue context to improve the accuracy of spatial transcriptomics predictions from H&E images. This proved beneficial across diverse datasets (Figure 2A, S1). In contrast, the linear regression and MLP baseline performed comparably to the previous models, ST-Net and BLEEP, which use only the target spot as input. Moreover, BLEEP inference requires complete access to the training data for k-nearest neighbor mapping in the latent space, which may become impractical due to the increasing data size and various ethical and privacy requirements in clinical setting. Furthermore, the vision transformer models struggled to establish robust correlations between morphology and transcriptomics possibly due to their inability to capture the spatial tissue environment and their limitations in incorporating morphological features from pre-trained image models.

Several promising paths exist to enhance DeepSpot’s gene expression prediction. 1) Currently, we split each spot into a fixed number of non-overlapping sub-spots, but this approach could be improved by using a cell segmentation algorithm to detect cell locations and boundaries, enabling us to divide the spot based on identified cells. 2) While we utilized general pathology foundation models to extract tile representations, it would be beneficial to use models specifically tailored to particular tissue types or tasks that are more relevant to spatial transcriptomics prediction (e.g., Hover-Net [63], cell nuclei segmentation and classification). Additionally, we anticipate that advancements in pathology foundation models could further enhance DeepSpot’s performance. 3) Currently, all neighboring spots are assigned the same weight; however, as the neighboring radius increases, this assumption may not hold. Therefore, it is important to investigate how to account for this variability such as by setting fixed radius parameters or learning a neighboring weight function. 4) The single-cell model used nuclei locations from Xenium ground truth data. In practical applications, these must be inferred computationally, requiring accurate nucleus and cell detection. Improving these methods and integrating them into the pipeline will be essential for enabling singlecell resolution in real-world settings.

In the future, DeepSpot could be applied to accurately predict spatial transcriptomics from H&E images and generate large-scale multimodal spatial transcriptomics datasets. This advancement would make spatial transcriptomics more accessible to scientists and healthcare professionals, enhancing its potential for biological discovery and accelerating clinical adoption. To foster these efforts, we have released the code for DeepSpot along with examples demonstrating its usage. Furthermore, we anticipate that the TCGA virtual spatial transcriptomics dataset, containing over 56 million spots, will serve as a valuable resource providing unique insights into the molecular landscapes of cancer tissues. It will also establish a benchmark for evaluating the performance and explainability of spatial transcriptomics models and support new model development. We hope that DeepSpot and this dataset will stimulate further advancements in computational spatial transcriptomics analysis.

## 4 Methods & Materials

### 4.1 Transcriptomics preprocessing

We normalized the transcriptomics count for each spot by total counts over all genes and then scaled it to a factor of 10,000, followed by a log1p transformation. To identify the most variable genes, we applied the *highly variable genes* function using Scanpy [34] with the Seurat v3 flavor, considering all spots in each dataset and using a batch key based on the tissue slice to reduce the impact of slice-specific variability. From this, we selected the top 5,000 most variable genes per dataset, which were used as the targets for prediction.

#### 4.1.1 Spatial oversampling with AESTETIK

Due to the skewed nature of the transcriptomics data, where genes might be expressed only in a small number of spots, we perform oversampling on the training data as follows: We utilize AESTETIK [5], a recent deep-learning model for spatial transcriptomics representation learning, to jointly integrate the spatial and transcriptomics modalities and project them into a lower-dimensional space for each slide. We then perform clustering in AESTETIK’s latent space using the Leiden algorithm with a resolution of 1 to obtain spot labels. Each cluster is subsequently oversampled to match the size of the largest cluster using resampling with replacement.

### 4.2 DeepSpot design

DeepSpot utilizes a deep-learning model based on deep-set neural networks [30]. The *φ*_*spot*_ module consists of a single fully connected layer with dropout regularization and ReLU activation. Its weights are shared across the three submodules—spot, sub-spot, and neighboring spots—enabling the model to perform multiple tasks simultaneously: 1) extracting features specific to each spot, 2) aggregating information from sub-spot features, and 3) maximum pooling features across the neighboring spots. The outputs from these submodules are concatenated and used as inputs to the *ρ*_*gene*_ module, which also consists of a single fully connected layer with dropout regularization and ReLU activation. The *ρ*_*gene*_ module predicts gene expression in a multiregression setting. To enhance network stability, we employ an ensemble architecture for both *φ*_*spot*_ and *ρ*_*gene*_ modules using random LeCun initialization [64]. The ensemble output is obtained by averaging the predictions from an ensemble of 10 *φ*_*spot*_ and 10 *ρ*_*gene*_ modules. DeepSpot is implemented in Python using PyTorch and PyTorch Lightning.

#### 4.2.1 Model input

For each spot tile *X* with array coordinates *x array*_*target*_ and *y array*_*target*_, we compute 3×3 nonoverlapping sub-spot tiles and select the closest neighboring spot tiles within a radius *r* from the target spot’s location in the array (*x array*_*target*_−*r <*= *x array <*= *x array*_*target*_+*r*; *y array*_*target*_−?*r <*= *y array <*= *y array*_*target*_ + *r*). For each tile, we extract spot features using recent pathology foundation models (e.g., UNI [20], Phikon [21], H-optimus-0 [31]), which were pre-trained on extensive H&E datasets, and follow their recommended image preprocessing workflows. For each spot *X*, we construct three sets: one representing the spot itself, one representing the local tissue structure (3×3 - 9 sub-spots), and one representing the global tissue environment (neighboring spots located within a radius *r*).

#### 4.2.2 Model output

We apply standard normalization per gene during training, using the mean and standard deviation computed from the training set. This process ensures that all genes are on the same scale, preventing the loss function from being dominated by a small subset of genes (Figure S1). In inference mode, we reverse the data transformation to restore the original gene ranges outlined in 4.1.

#### 4.2.3 Training details

The model is optimized using the mean squared error loss function (Equation 1). It is trained on a single GPU with at least 12GB RAM with early stopping, using the Adam optimizer [65] with a learning rate of 1e-4, a weight decay of 1e-6 and a batch size of 1024. Computational data analysis was performed at Leonhard Med (https://sis.id.ethz.ch/services/sensitiveresearchdata/), a secure trusted research environment at ETH Zurich.

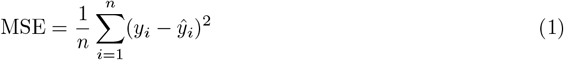

### 4.3 Evaluation

We evaluate the model’s performance using Pearson correlation per gene (Equation 2), employing nested leave-one-out patient cross-validation with an internal 3-fold cross-validation loop for hyperparameter selection. Specifically, for each fold, we trained a model on the training set, selected hyperparameters on the validation set, and evaluated performance on the test set, ensuring that each set contained distinct patients. This process was repeated across all folds. We bootstrapped 10,000 times from the median Pearson correlation across the test folds and reported the resulting median Pearson correlation along with its standard error. For state-of-the-art methods, we utilized hyperparameter values recommended by the authors or discussed in their respective papers. The specific hyperparameter settings are included in the supplementary materials.

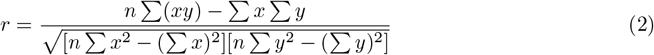

#### 4.3.1 Out-of-distribution evaluation on TCGA

Moreover, for out-of-distribution evaluation on TCGA, we generated spatial transcriptomics samples from the available TCGA image slides (FFPE and FF) and applied the pre-trained DeepSpot model (trained on the full spatial transcriptomics dataset for the specific condition) to predict gene expression. We then computed pseudo-bulk expression by averaging gene expression across all spots per slide and measured the correlation with the corresponding bulk RNA-seq data from TCGA. While the FF slides and their corresponding bulk RNA profiles came from the exact same tissue piece, the FFPE slides with larger tissue areas lacked exact bulk RNA profiles. Therefore, we matched them to a bulk RNA profile based on patient-level identification and tumor type classification. We applied the pre-trained melanoma model to TCGA samples with skin melanoma (TCGA SKCM, n=472 FF; n=276 FFPE), the kidney cancer to TCGA samples with renal cell (TCGA KIRC, n=528 FF; n=516 FFPE) and the lung cancer [26] to TCGA samples with lung adenocarcinoma (LUAD, n=537 FF; n=525 FFPE) or lung squamous cell carcinoma (LUSC, n=455 FF; n=471 FFPE).

#### 4.3.2 Ablation study

In the ablation study of DeepSpot, we follow the procedure outlined in 4.3, where we fix the hyperparameter of interest and evaluate its effect on the Area under the Pearson gene correlation curve (Figure S1). We studied the impact of the neighbor spot radius, loss function, feature image model, normalization and oversampling.

### 4.4 Downstream applications

#### 4.4.1 Predicted gene expression transformation

We enforce positive gene values by setting all values smaller than 0 to 0 and adding 1. This approach ensures compatibility with downstream tasks without introducing additional computational challenges (e.g., NaN values due to negative scores or floating point overflow) or requiring custom solutions.

#### 4.4.2 Marker genes

For identifying marker genes, we use the *rank genes groups* function from Scanpy with the Wilcoxon signed-rank test. We select significant marker genes and rank them by their average log-fold change.

#### 4.4.3 Gene set enrichment analysis

For the gene set enrichment analysis, we utilized the Python interface for EnrichR [66] in conjunction with data from the Cancer Cell Line Encyclopedia [45]. We selected the top 100 tumor marker genes based on their log-fold change.

#### 4.4.4 Pathway analysis

For pathway analysis, we apply the multivariate linear model from the *decoupler* package [51] to assess regulatory pathway activities, using data from the *PROGENy* database [67].

#### 4.4.5 Gene Ontology enrichment analysis

We identified the top 10% of genes per dataset based on their Pearson correlation with the ground truth and conducted gene ontology enrichment analysis using *EnrichR* [68] and *GO Biological Process 2025* terms [69]. Significant pathways were selected based on an adjusted p-value ¡ 0.05 and their presence in at least 2 datasets. To further refine the results, we employed *simplifyEnrichment* [70] to compute pathway similarity and identify common terms. For low-ranking genes, we selected those in the bottom 50th percentile per dataset, supported in at least 3 of 5 datasets, to capture weaker individual signals that may indicate consistent biological relevance.

#### 4.4.6 Survival analysis on TCGA

We used the Cox proportional hazards model with an elastic net penalty [71] to conduct the survival analysis benchmark to predict the overall survival as defined in [72]. For all experiments, we performed outer 5-fold cross-validation, repeated 10 times and stratified on the event indicator. We reported the mean concordance index along with its standard error. Data splits were performed using the *StratifiedKFold* implementation from *scikit-learn*. To ensure comparability across slides, we randomly selected 128 spots from each slide with replacement and averaged their corresponding gene expression values. We selected the one with the largest tissue area when multiple samples were available per patient.

#### 4.4.7 Tumor type classification on TCGA

Following the procedure described in 4.4.6, we implemented logistic regression with an L2 penalty and evaluated the model using the F1 score.

#### 4.4.8 Super-resolution gene expression prediction

To obtain super-resolution gene expression predictions, we utilize the already pre-trained DeepSpot. Rather than feeding all non-overlapping sub-spots into the model, we select only a sub-spot in the center and adjust the neighbor spot distances accordingly.

#### 4.4.9 H&E images and single-cell atlas integration

To integrate the H&E images, we predicted the spatial transcriptomics profiles using the H&E images. We then aligned the predicted profiles with the human lung reference atlas [62] in the PCA latent space computed using the genes shared between the datasets. Missing gene expression was inferred through linear projection by fitting a regression model on the reference atlas to predict the unobserved genes. For annotation transfer, we employed the *ingest* function in Scanpy.

### 4.5 Spatial trancsriptomics data

We used spatial transcriptomics datasets generated using Visium from 10x Genomics™ with a capture area of 6.5×6.5mm and a spot diameter of 55 μm, as well as datasets from 10x Genomics™ Xenium, which offers subcellular resolution.

#### 4.5.1 Tumor Profiler

The Tumor Profiler dataset [5] consists of 18 tissue slices from 7 patients with metastatic melanoma and includes pathology annotations with the following labels: Tumor, Normal lymphoid tissue, Blood-/Necrosis, Stroma, and Pigment. The dataset consists of FFPE sections processed using Visium from 10x Genomics™ technology. The H&E slices and corresponding pixel coordinates were scaled to 20x magnification.

#### 4.5.2 Kidney, Colon and Lung cancer HEST-1K

The kidney cancer [28] dataset consists of 24 tissue slices from 24 patients with kidney cancer. The colon cancer [29] consists of 8 tissue slices from 4 patients. Both datasets consist of fresh frozen sections processed using Visium technology and were downloaded from the HEST-1K [14]. Although the H&E slices were originally available at 40x magnification, to ensure compatibility with the TCGA slides, which are a mixture of 20x and 40x, we scaled the H&E slices and corresponding pixel coordinates to 20x magnification. The lung cancer dataset [27] consists of 20 tissue slices from 19 patients with lung cancer. It consists of FFPE sections processed using Xenium from 10x Genomics™ technology.

#### 4.5.3 Kidney and Lung cancer with TLS

The kidney and lung cancer with tertiary lymphoid structures (TLS) dataset consists of 5 *μ*m thick FFPE sections from kidney (3) and lung (5) tumors obtained from the Institute of Pathology at the University Hospital of Zurich and mounted onto the Visium slides with the Human Probe Set v1. Samples were stained with hematoxylin and eosin and subsequently processed for sequencing following the manufacturer’s recommendations. After library preparation, the samples were sequenced on an Illumina Novaseq 6000 and preprocessed with Space Ranger v2.1.0. Patient analyses were conducted according to the Declaration of Helsinki. Ethical approval for performing research on anonymized, archival patient material was obtained from the cantonal ethics commission Zurich (BASEC Nr. 2022-01854 and BASEC-Nr. 2024-01428). Spatial transcriptomics sequencing was performed at the Functional Genomics Center Zurich (FGCZ) of the University of Zurich and ETH Zurich. The Visium spots were annotated in the corresponding H&E images by expert researchers (K.S. and S.D.) and included manual annotations with the following labels: TLS (tertiary lymphoid structures), Immune, Tumor, Normal, Lymph nodes and Unassigned. One lung (LC4) and one kidney (KC2) slice were excluded from the analysis due to a misalignment between the spatial transcriptomics data and the expected morphological features (Figure S5).

#### 4.5.4 Lung cancer

The lung cancer dataset comprises 36 tissue slices from 8 patients diagnosed with non-small cell lung cancer [26]. Each patient contributes 4 FFPE slides, each containing two tumor regions and two background (non-tumor) regions. The dataset consists of FFPE sections processed using Visium from 10x Genomics™ technology. The H&E slices were available at 20x magnification.

#### 4.5.5 TCGA

For out-of-distribution validation and de novo spatial transcriptomics prediction, we used patient metadata, image slides (fresh frozen - FF and formalin-fixed paraffin-embedded - FFPE) and paired bulk RNA-seq from the TCGA Research Network (https://www.cancer.gov/ccg/research/genome-sequencing/tcga) from samples with skin melanoma (SKCM), renal cell (KIRC), lung adenocarcinoma (LUAD) and lung squamous cell carcinoma (LUSC). When multiple FF sections were available for the same tissue sample, the top section was selected. All slides were scaled to 20x magnification. To distinguish between in-tissue spots and background, we computed the mean RGB value for each tile and discarded all tiles with a mean greater than 200 (close to white).

## 4.6 Data availability

The kidney, colon and lung cancer HEST-1K datasets were downloaded using the HEST-1K download pipeline outlined here: https://github.com/mahmoodlab/HEST/blob/main/tutorials/1-Downloading-HEST-1k.ipynb, with data available at Hugging Face https://huggingface.co/datasets/MahmoodLab/hes. The Tumor Profiler spatial transcriptomics dataset was obtained from [5]. The lung and kidney cancer with TLS dataset is available at https://zenodo.org/records/14620362. The human lung reference atlas is available at https://data.humancellatlas.org/hca-bio-networks/lung/atlases/lung-v1-0. The lung cancer dataset was downloaded from https://www.ebi.ac.uk/biostudies/arrayexpress/studies/E-MTAB-13530. The single-cell lung reference atlas was downloaded from https://data.humancellatlas.org.

TCGA virtual spatial transcriptomics samples are available for download on Hugging Face. Access details can be found at https://github.com/ratschlab/DeepSpot.

## 4.7 Code availability

The open-source implementation of DeepSpot, along with a tutorial is available at:

- https://github.com/ratschlab/DeepSpot

The Snakemake pipeline for reproducing the results is available at:

- https://github.com/ratschlab/he2st

## Acknowledgments

This work was supported by the Swiss Federal Institutes of Technology (strategic focus area of personalized health and related technologies; 2021–367). The 10x spatial transcriptomics sequencing of a subset of the Tumor Profiler samples was made possible through a technology access program by 10x Genomics™, with special acknowledgments to Jacob Stern, James Chell, Rudi Schläfli, Laura Lipka, Mario Werner, Nikhil Rao, and Scott Brouilette for their invaluable contributions. The Tumor Profiler study was supported by a public-private partnership involving Roche Holding AG, ETH Zurich, University of Zurich, University Hospital Zurich, and University Hospital Basel.

## Consent for publication

This manuscript has been seen and approved by all listed authors. The figures were created using BioRender.com and exported under a paid subscription.

## TUMOR PROFILER CONSORTIUM

