## Supplementary information for "DeepSpot: Leveraging Spatial Context for Enhanced Spatial Transcriptomics Prediction from H&E Images"

#### 1 Comparison with other methods

For comparing other spatial transcriptomics methods, we applied the suggested preprocessing pipelines outlined in the corresponding papers and tutorials. The considered hyperparameter values suggested by the authors, as well as those discussed in the corresponding paper can be found in table S1. MeanCellType, Linear Regression and Multilayer perceptron (MLP) serve as our baseline models, using the same pathology foundation models to extract features while utilizing only the target spot. MeanCellType applies the Leiden algorithm to Harmony-integrated [1] data to define cell types, then trains an MLP to predict these types and estimates gene expression profiles by averaging expressions within each cell type in the training set. Similar to [2], for the vision transformer-based model, we randomly selected 128 spots per slide to ensure it fits into GPU memory.

| Model | Hyperparameters |
|---|---|
| *MeanCellType* | image model: $[phikon, uni, hoptimus0]$; resolution: $[0.1, 0.5, 1, 10]$ |
| *LinearRegression* | image model: $[phikon, uni, hoptimus0]$ |
| *MLP* | image model: $[phikon, uni, hoptimus0]$; hidden_layer_sizes: $[[256, 256], [512, 512]]$ |
| *DeepSpot* | image model: $[phikon, uni, hoptimus0]$; oversampling: $[True, False]$; gene_norm: $[none, standard]$ |
| *STNet [3]* | image model: $[densenet121]$; learning_rate_init: $[0.00001, 0.000001]$; batch_size: $[32, 64]$; solver: $[SGD]$; momentum: $[0.9]$ |
| *BLEEP [2]* | image model: $[resnet50]$; epochs: $[150]$; batch size: $[512]$; top k: $[1, 50, 100]$ |
| *HisToGene [4]* | dropout: $[0.2, 0.4]$; n layers: $[4, 8]$; n pos: $[128]$; batch size: $[1]$, epochs: $[100]$ |
| *Hist2ST [5]* | dropout: $[0.2, 0.4]$; heads: $[8, 16]$; n pos: $[128]$; batch size: $[1]$, epochs: $[350]$ |
| *THItoGene [6]* | dropout: $[0.2, 0.4]$; n layers: $[4, 8]$; n pos: $[128]$; batch size: $[1]$, epochs: $[350]$ |

**Table S1**: Model hyperparameters.

### 2 Suplementary figures

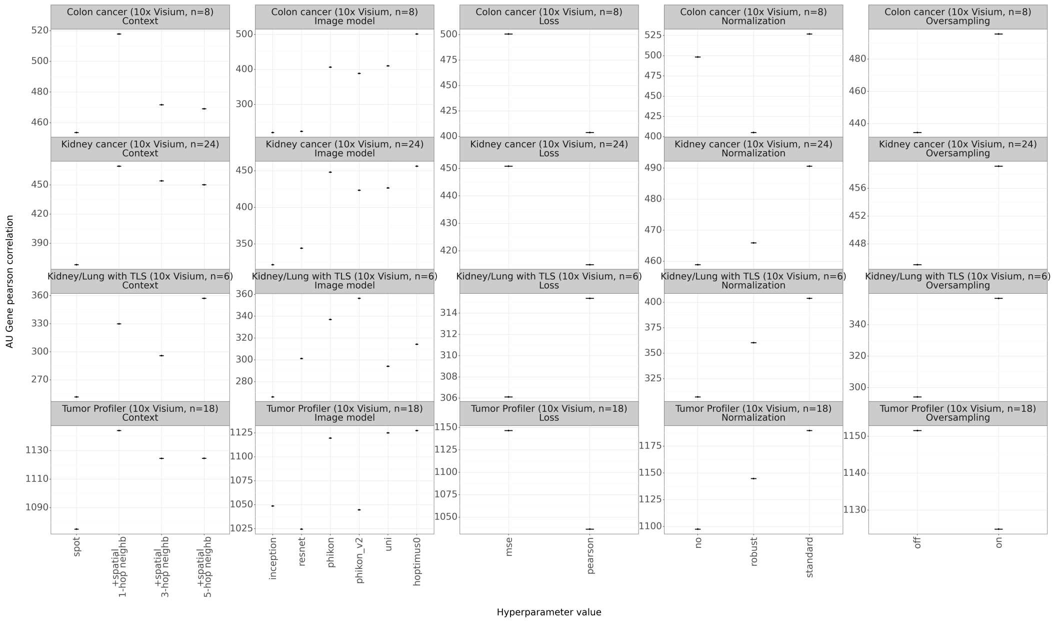


**Fig. S1**: **Ablation study on DeepSpot's hyperparameters.**
The Y-axis represents the area under the Pearson correlation curve for the most predictive gene subsets, while the X-axis shows the various hyperparameter values.

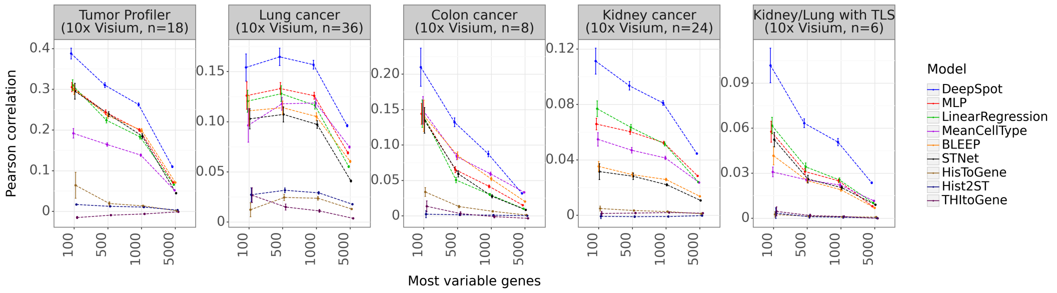


**Fig. S2**: **Most variable genes benchmark of DeepSpot and previous state-of-the-art methods across multiple spatial transcriptomics datasets generated using Visium from 10x Genomics™**
The y-axis represents the average Pearson correlation between the predicted and ground truth gene expression. The x-axis represents Pearson correlation computed on the top N highly variable genes as defined by Scanpy with the Seurat v3 flavor [7] on the training data. Models are ordered based on their relative rank across datasets.

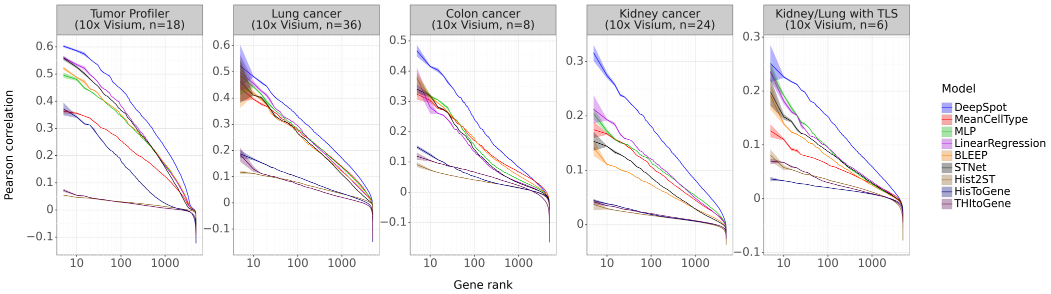


**Fig. S3**: **Gene rank benchmark of DeepSpot and previous state-of-the-art methods on spatial transcriptomics datasets generated using Visium from 10x Genomics™**
The y-axis represents the average Pearson correlation between the predicted and ground truth gene expression for each gene rank (x-axis) per model, where rank 1 indicates the best and 5000 the worst. Models are ordered based on their relative rank across datasets.

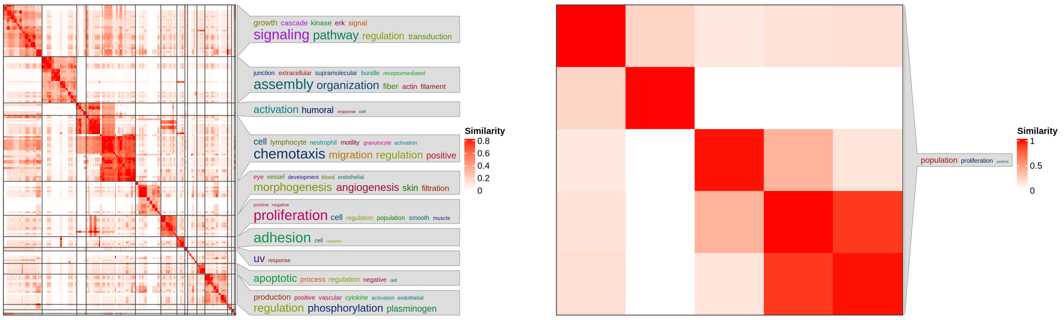


(a) Pathways associated with well-predicted genes, defined as those in the top 10th percentile per dataset and supported by at least 2 out of 5 datasets.

(b) Pathways associated with lower-ranked genes, defined as those in the lower 50th percentile per dataset and supported by at least 3 out of 5 datasets.

**Fig. S4**: **Simplify Gene Ontology (GO) enrichment results.**
Pathway enrichment analysis was performed using EnrichR in conjunction with the GO Biological Process 2025 database [8], followed by identification of common biological processes through simplifyGO using default parameters [9]. The plot illustrates the semantic similarity between GO terms, organized into functional clusters.

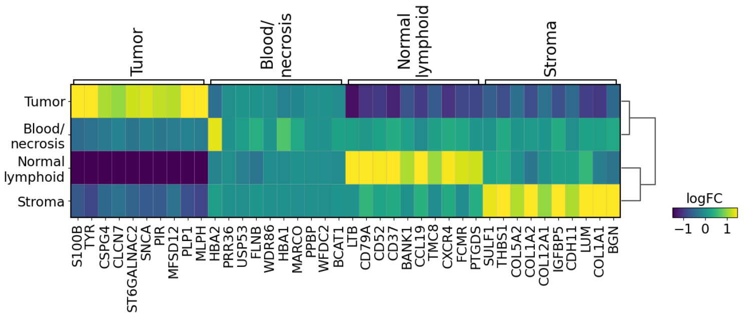


(a) Virtual spatial transcriptomics generated by DeepSpot.

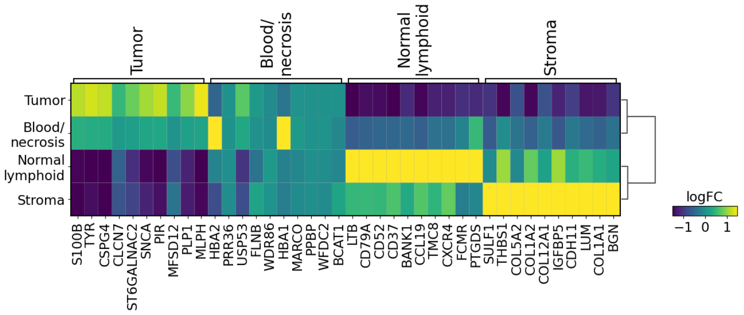


(b) Ground truth spatial transcriptomics generated by Visium from 10x Genomics.

**Fig. S5**: Comparison of matrix plots showing the log fold change (logFC) of the top 10 marker genes per group for sample MELIPIT-1-2.

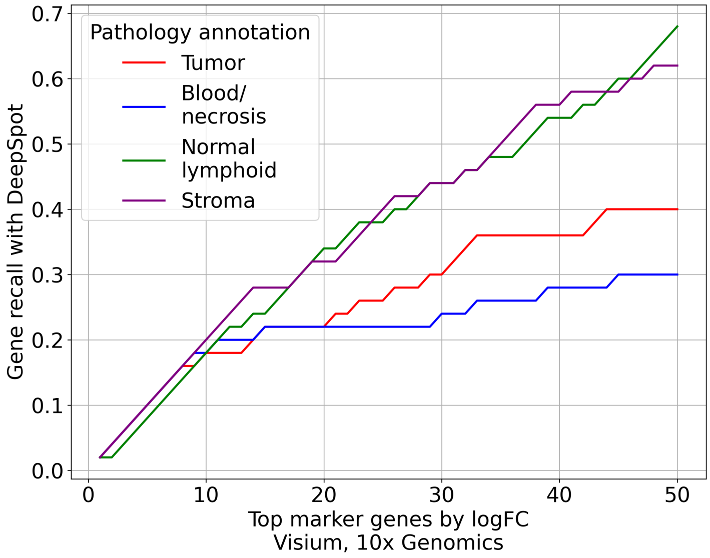


**Fig. S6**: **Recall of the significant top 50 marker genes per group.**
The top 50 significant marker genes per group, ranked by log-fold change (logFC), were identified using the ground truth spatial transcriptomics data and compared to those selected by DeepSpot using the same criteria. The stroma and normal lymphoid groups achieved a recall of almost 0.7, whereas the tumor group reached a recall of 0.4.

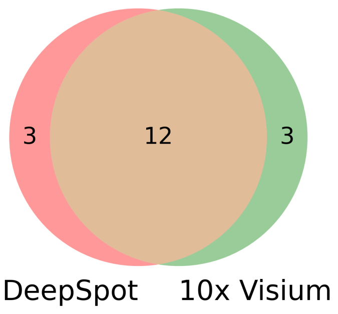


**Fig. S7**: **Overlap of the top 15 cell lines identified using ground truth spatial transcriptomics data and virtual spatial transcriptomics.**
The two methods showed an overlap of 12 out of 15 pathways, corresponding to 80%.

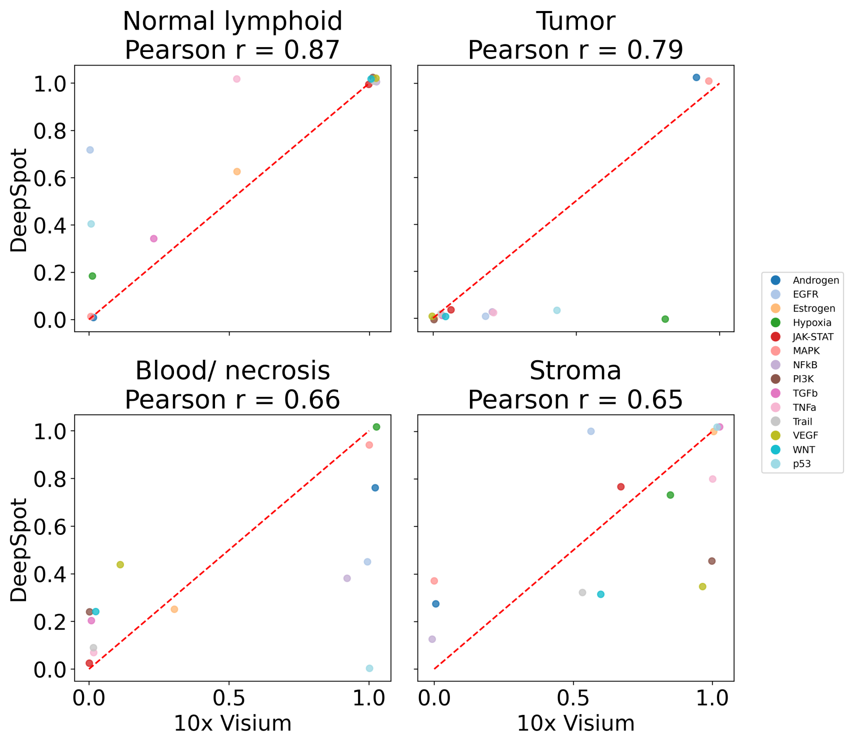


**Fig. S8**: **Pathway activity correlation between ground truth spatial transcriptomics and virtual spatial transcriptomics.**
Pathway activity scores were independently computed for both ground truth and virtual spatial transcriptomics datasets using decoupleR [10]. The similarity between the two was then quantified by calculating the Pearson correlation of the pathway activity scores.

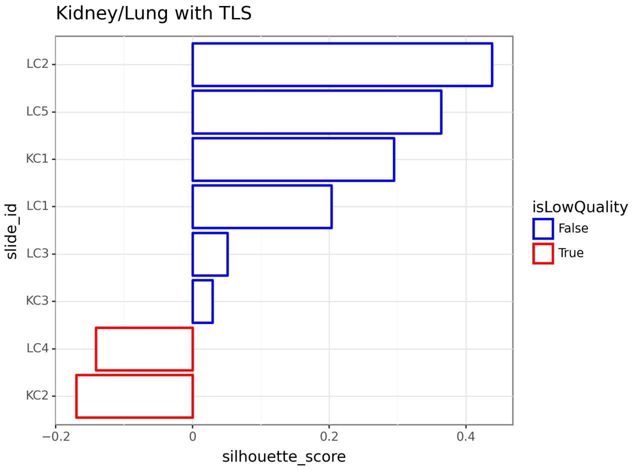


**Fig. S9**: **Alignment of the gene expression with the manual annotations.**
Silhouette score calculated using the first two principal components derived from PCA on the log-normalized gene expression data and the provided manual annotations.

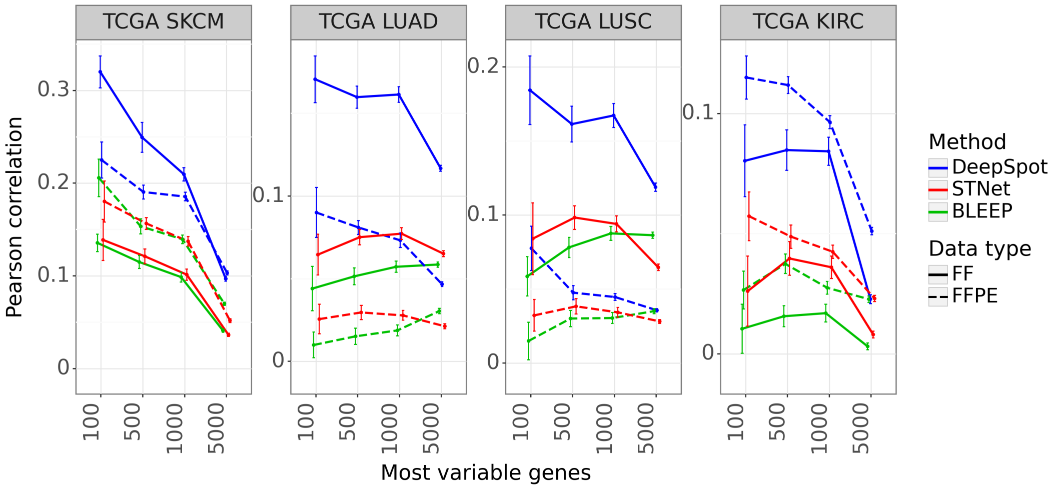


**Fig. S10**: **Most variable genes out-of-distribution benchmark on models trained on 10x Genomics™, Visium and gene expression predictions based on TCGA slides.**
The y-axis represents the Pearson correlation between the pseudo-bulk RNA and ground truth bulk RNA profile. The x-axis represents Pearson correlation computed on the top N highly variable genes as defined by Scanpy with the Seurat v3 flavor [7] on the training data.

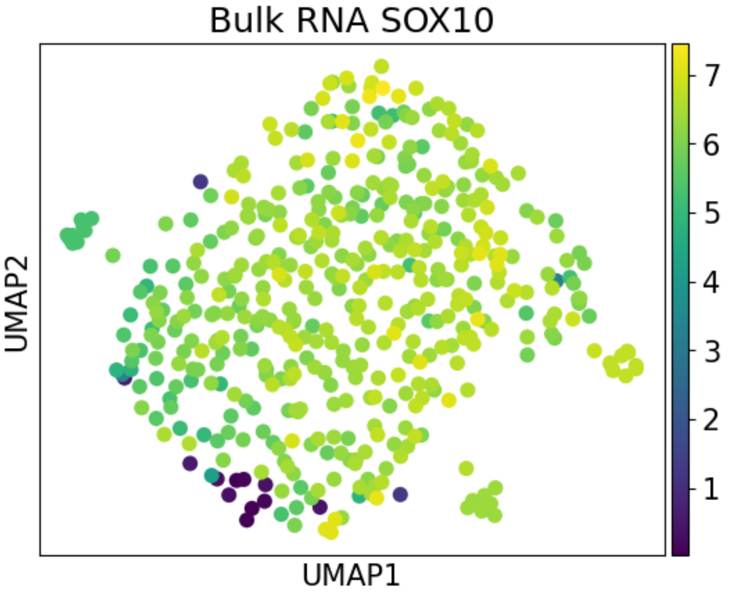


**Fig. S11**: **UMAP of the ground truth TCGA SKCM bulk RNA-seq, colored by *SOX10* gene expression.**

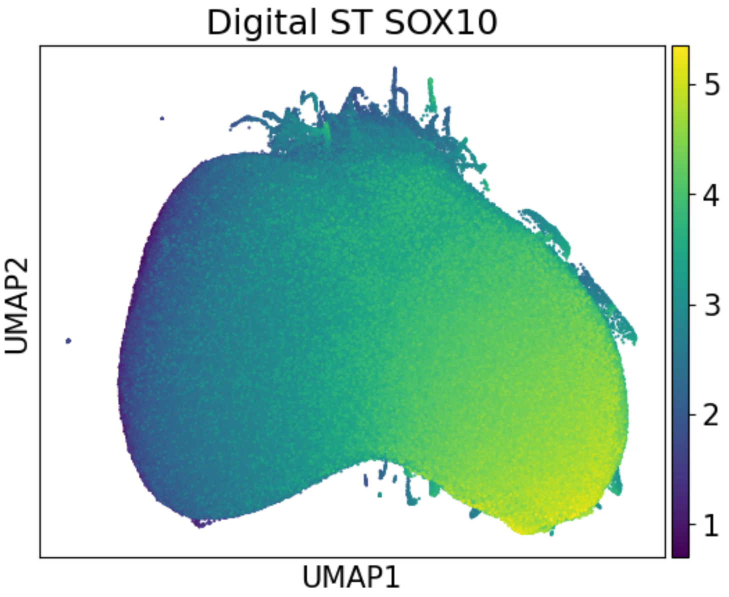


**Fig. S12**: **UMAP of the predicted spatial transcriptomics from TCGA SKCM image slides, colored by *SOX10* gene expression.**

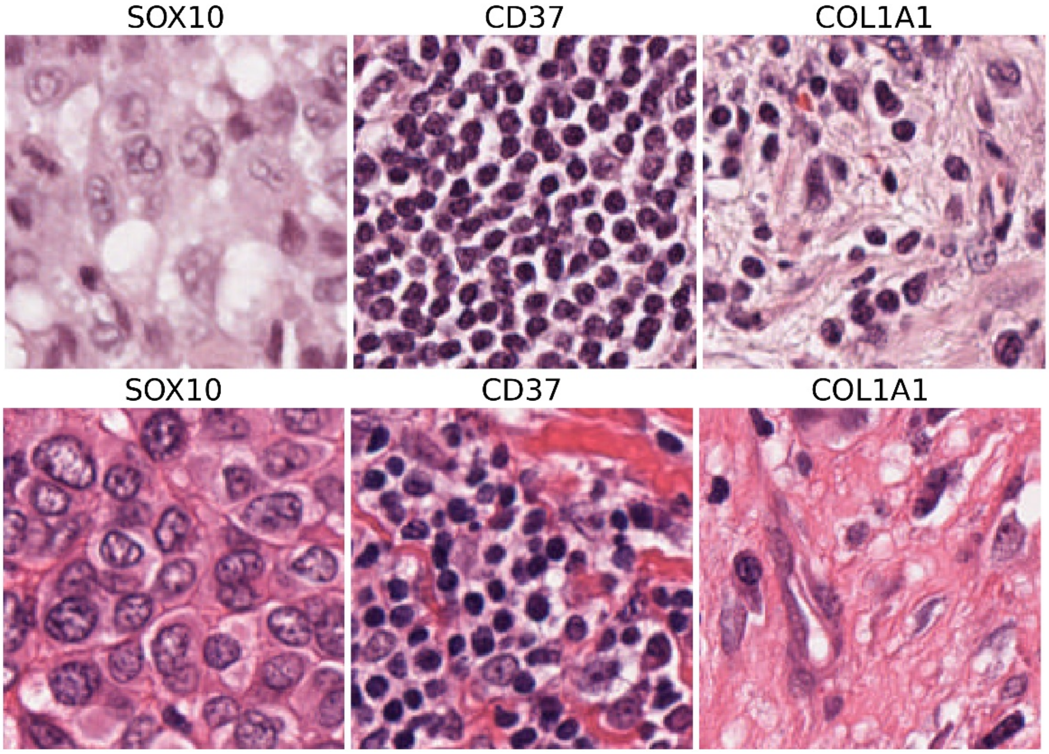


**Fig. S13**: **TCGA FFPE SKCM examples of spot images for *SOX10* (melanoma), *CD37* (normal lymphoid), and *COL1A1* (stroma).**

### 3 Suplementary tables

| Hold-out-genes | | | | | | | |
|---|---|---|---|---|---|---|---|
| HEY1 | ACKR1 | CA4 | FCN3 | APLNR | COL1A1 | POSTN | CTHRC1 |
| MSLN | WNT5A | ACTA2 | MFAP5 | CSPG4 | PTGDS | PLIN2 | |

**Table S2**: List of hold-out-genes used for validation in the Xenium experiment in Figure 6C and 6G.
